# COVID-19-associated school closures and related efforts to sustain education and subsidized meal programs, United States, February 18–June 30, 2020

**DOI:** 10.1101/2021.03.05.21252848

**Authors:** Nicole Zviedrite, Jeffrey D. Hodis, Ferdous Jahan, Hongjiang Gao, Amra Uzicanin

## Abstract

Pre-emptive school closures are frontline community mitigation measures recommended by CDC for implementation during severe pandemics. This study describes the spatiotemporal patterns of publicly announced school closures implemented in response to the coronavirus disease 2019 (COVID-19) pandemic and assesses how public K-12 districts adjusted their methods of education delivery and provision of subsidized meals. During February 18–June 30, 2020, we used daily systematic media searches to identify publicly announced coronavirus disease 2019 (COVID-19)–related school closures lasting ≥1 day in the United States (US). We also collected statewide school closure policies from state government websites. Data on distance learning and subsidized meal programs were collected from a stratified sample of 600 school districts. The first COVID-19–associated school closure occurred on February 27, 2020 in Washington state. By March 30, 2020, all but one US public school districts were closed, representing the first-ever nearly synchronous nationwide closure of public K-12 schools in the US. Approximately 100,000 public schools were closed for ≥8 weeks because of COVID-19, affecting >50 million K-12 students. Of 600 districts sampled, the vast majority offered distance learning (91.0%) and continued provision of subsidized meal programs (78.8%) during the closures. Despite the sudden and prolonged nature of COVID-19–associated school closures, schools demonstrated flexibility by implementing distance learning and alternate methods to continue subsidized meal programs.

## Introduction

Preemptive school closures are implemented before the transmission of respiratory infectious disease, such as influenza, is widespread in schools and surrounding communities. They are recognized as one of the most impactful nonpharmaceutical interventions (NPIs) used to slow the spread of influenza pandemics [1,2].

As of the summer 2020, the severe acute respiratory syndrome coronavirus 2 (SARS-CoV-2), the causative agent of coronavirus disease 2019 (COVID-19), had caused the world’s most severe pandemic since the 1918 influenza pandemic [3]. The first cases, characterized as pneumonia of unknown etiology, were reported in Wuhan, China in December 2019, and were soon followed by spread into, within, and between other countries [4]. On January 21, 2020, the first case was reported in the United States (US) [5]. The World Health Organization declared COVID-19 a pandemic on March 11, 2020 [6]. The emergence of COVID-19 has led to unprecedented use of NPIs worldwide, including prolonged SCs, in order to slow the disease spread.

Following the first identified case in the US, an additional 13 cases were identified during the weeks leading up to February 23^rd^ but were limited to persons with recent travel from China and their household contacts [7]. In response, the Centers for Disease Control and Prevention (CDC) first posted interim guidance for K-12 schools in mid-February, which provided guidelines for closing schools with recognized cases of COVID-19 among students, staff, or visitors [8].

In late February, the first known US cases of COVID-19 in persons who had no history of recent travel to countries with ongoing community transmission were reported. By mid-March, all 50 states, the District of Columbia (DC), and all four US territories had reported at least one case of COVID-19 [9]. To protect healthcare capacity and slow community spread of COVID-19, various local, state, and federal authorities issued stay-at-home orders and the closing of some schools and nonessential workplaces [7].

SCs present the challenge of providing continuity of education and other school-based services, such as subsidized meal programs. The objectives of this study are to examine both the extent of COVID-19–associated K-12 SCs in the US during the initial five months, February–June 2020, of the COVID-19 pandemic, and to describe how the affected schools responded to the disruption with strategies for the continued provision of education and subsidized meals programs throughout closure.

## Methods

Using previously described methodology [10], we conducted daily searches of publicly available online data (via Google, Google News, Google Alerts). Searches were conducted using the following terms in Google search and Google News, “school closed” and “COVID,” “COVID-19,” or “coronavirus”. Searches in predefined Google Alerts used the following search string, “(academy OR school OR district OR class) AND (close OR closing OR closure OR cancel OR cancelled) AND (coronavirus OR corona OR “COVID-19” OR COVID OR “novel coronavirus”)”. These searches derived both national and local news media sources, as well as government, district, and school websites that shared public announcements of illness-related unplanned closures of individual public schools and districts lasting at least one day in the US during February 18– June 30, 2020. We selected SCs for which COVID-19 was given in public announcements as the reason for the closure (hereafter referred to as COVID-SCs).

Additionally, with the announcement of the first statewide closures on March 12, 2020, we augmented our search strategy by performing daily searches of Google (combining initial search terms with individual state names) and state government websites (manually identifying and searching the websites of state governors, and health and education authorities) in order to identify and track state-level mandates and recommendations for COVID-SCs from governors and/or state education or health authorities. For statewide closure mandates, we populated our database with all K-12 school districts, which are by definition public, in the respective states using the effective date of the mandate as the date of closure unless the district separately announced closure before the mandate went into effect. All school districts in the National Center for Education Statistics (NCES) Common Core of Data, including regular school districts, independent charter districts, etc., with one or more schools and one or more students were included [11]. For statewide closure recommendations, we looked for confirmation of compliance with the recommended policy through state government and media sources. In instances where compliance was unclear, we manually confirmed closures using the information found on school district websites. Because of the unprecedented level of closure, the capture of individual school closures was limited to during February 18–March 24, 2020, at which point statewide public COVID-SCs were in affect across the US.

From the NCES website, we downloaded publicly available school-related data for the most recent school year available (public schools and districts, 2017–2018; private schools, 2017-2018) [11]. We recorded data on school and school district characteristics including the number of schools in affected districts, the number of students, the percentage of students enrolled in the federal free or reduced-price school lunch program, and location information (latitude, longitude, county, and state). We linked these data to identified COVID-SCs using the NCES district or individual school identifiers.

NCES school districts were divided into quartiles based on the percentage of students eligible for free/reduced-price lunch, an indicator of family economic status [12]. A simple random sample was taken in each stratum, and sample size per stratum was calculated using 95% confidence interval of 50% ± 10%. The sample included school districts from all 10 HHS regions, 46 US states, and the District of Columbia (DC); the school districts in the sample ranged in size from 1–92 schools (1–63,223 students). After excluding one school district that per the NCES database had one school with one student, the range was nearly unchanged (1–92 schools and 3–63,223 students). We collected data on the availability and method of delivery of both distance learning and subsidized meal programs via public announcements on school district websites and their official social media pages (Facebook, Twitter). Google searches were performed for news resources when information was not available from official, online district sources.

Finally, we obtained publicly available online surveillance data on COVID-19 cases from the COVID-19 Data Repository by the Center for Systems Science and Engineering (CSSE) at Johns Hopkins University [13].

COVID-SC data were imported into SAS 9.4 (SAS Institute Inc., Cary, North Carolina) for analysis. Descriptive statistics were generated to summarize school characteristics and the availability of distance learning and subsidized meal programs during COVID-SCs. Maps illustrating public school district closures, the timing of statewide public school closure policies, and types of statewide policies for private school closures were generated using Microsoft Power BI.

## Results

### Individual and district level closures

The first known COVID-SC in the US was an individual SC on February 27, 2020 in Washington State, which was reported in response to a suspected COVID-19 case in the household of a school employee. By the following week (March 1–7, 2020), both public school- and district-level closures occurred in at least 11 states because of one or more confirmed or suspected cases in the school or district, a cluster in the community, or as a precautionary measure (Fig 1A). During that week, there were 15 district-wide and 33 individual public school COVID-SCs, for an estimated total of 240 closed public schools. During the following week, March 8–14, before any statewide SC went into effect, there were a total of 367 district closures and 91 individual COVID-SCs (Fig 1B). In response to growing reports of local transmission, increasing numbers of school districts began to close because of either local- or state-level SC decisions. Ultimately, more than 2,700 school districts (approximately 16% of the nation’s 16,000+ districts) closed prior to statewide closure orders going into effect in their respective states; these districts account for more than 18,000 schools and serve more than 16 million students.

**Figure 1.**
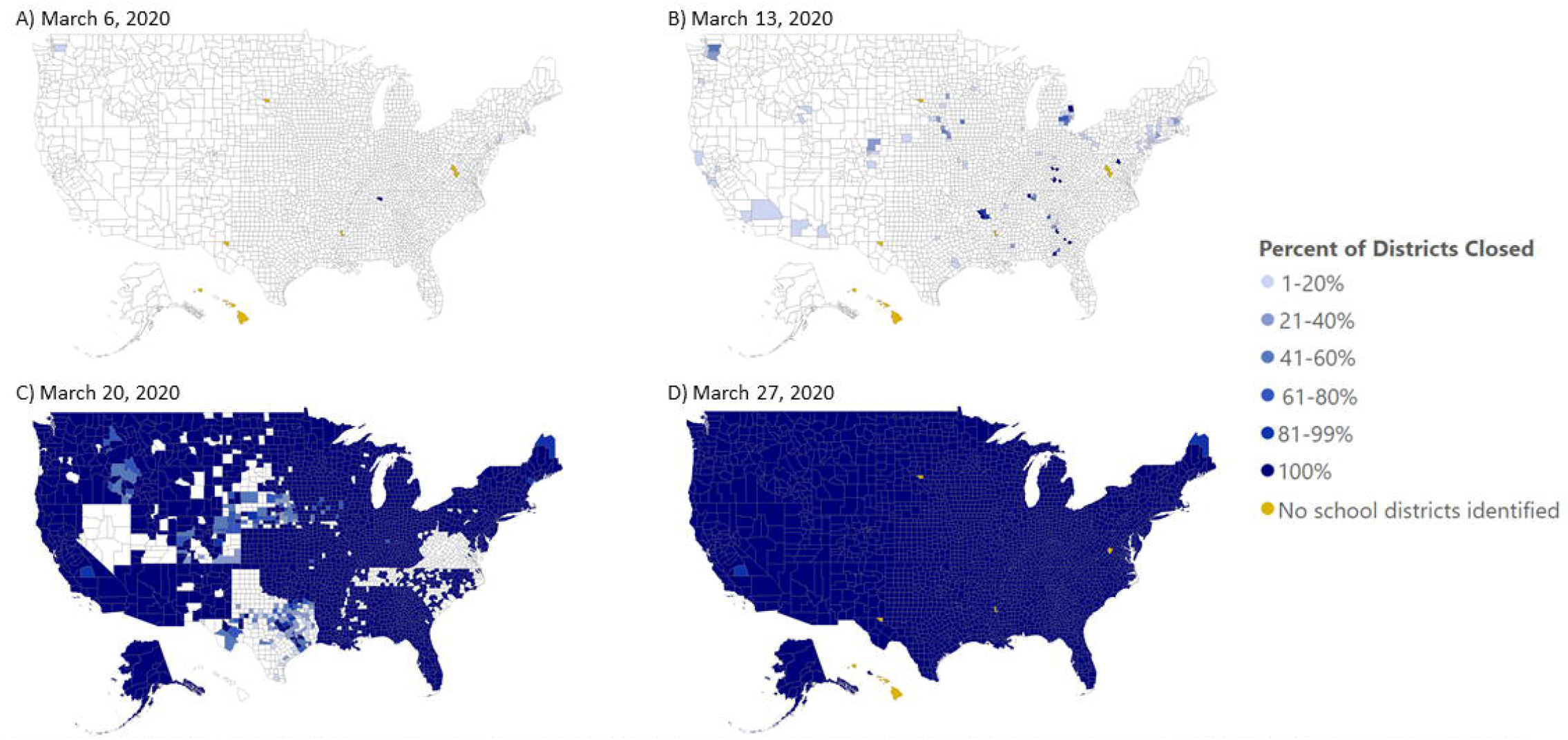
Coronavirus disease 2019-related Public K-12 School District Closures by County and Date - United States. **Data Sources, Reference, & Notes:** Data were collected via daily searches of publicly available content (Google Alerts, Google, and Google News). A standard set of search terms were used to identifyCOVID-1 related public school closures lasting ≥1 day. Official state-level health and education department websites, and school and district level websites were also searched. The map shows the county-level data on closures for the academic year 2019-2020for public K-12 school districts in which schools are physically closed to students for traditional on-site leaming. School districts may be using distance learning to continue education from home. Because data are limited to information derived from publicly available closure announcement some closures may have been missed depending on how they were reported and some information may not be complete or entirely accurate. Data are only collected for the 50 states and DC, are restricted public K-12 school districts, and do not include private schools. Number of districts per county was estimated per data available from the National Center for Education Statistics Common Core of Data (https.//nces.ed.gov/ccd/districtsearch/). Districts reporting 0 students were excluded. Yellow .shade denote the counties in which no public school districts have been identified. This is typically due to multiple counties being part of a single, consolidated school district, whereby the district is only reported in the country where the main office is located. Counties shown in white have no school district closures reported on that date.

### Statewide policies

Between mid- and late-March, statewide mandates or recommendations for public SCs were issued in every state, with the earliest announced in Kentucky, Maryland, Michigan, and Oregon on March 12, 2020. Among the 50 states and DC, 24 states and DC (49.0%) had public SCs go into effect on March 16. An additional 21 states had public SCs go into effect later that same week (March 17–21, 2020), and the remaining five states had public SCs go into effect during the week of March 22–28. Idaho was the last state to issue a mandated statewide public SC, which went into effect on Tuesday, March 24 (Fig 2, Table 1).

**Figure 2.**
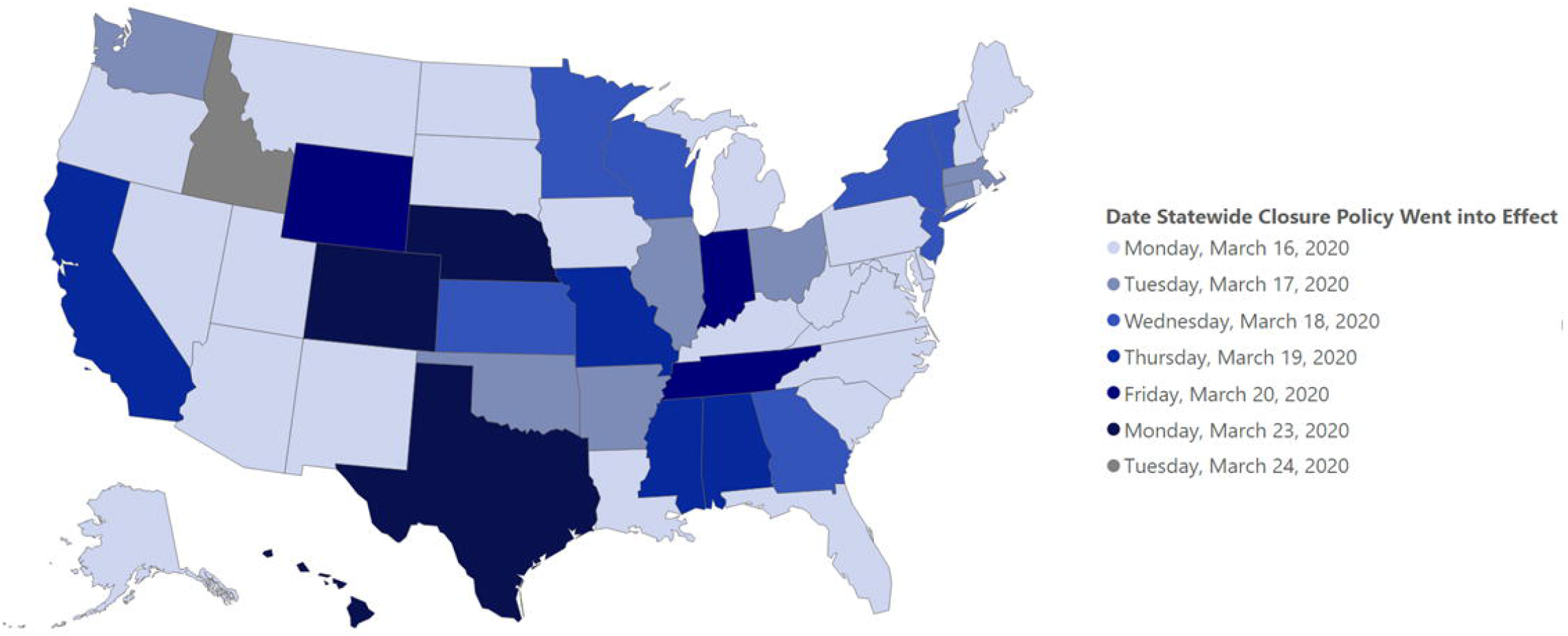
Effective Date of Statewide Coronavirus Disease 2019 Public K-12 School Closure Policies-United States, March 2020. **Data Sources, Reference, & Notes:** Data were collected via daily searches of publicly available online content (Google Alerts, Google, and Google News). A standard set of search terms were used to identify COVID-19-related public school closures lasting ≥1 day. Official state-level health and education department websites, and school and district-level websites were also searched. The map shows the effective date of state-level closure policies the academic year 2019-2020 for public K-12 school districts in which schools are physically closed for students. School districts may be using distance learning to continue education from home. Orders and recommendations for statewide closure originated from state governors and/or state education departments.

**Table 1.** Characteristics of statewide school and district closures during the coronavirus disease 2019 pandemic — United States, March 12–June 30, 2020.

| State | Type of policy | Issuer of order | Issue date | Effective date | End of school year status* | No. of public school districts | No. of public schools | No. of public school students |
| --- | --- | --- | --- | --- | --- | --- | --- | --- |
| Alabama | Mandate | Governor | March 13 | March 19 | CAY | 138 | 1,473 | 742,444 |
| Alaska | Mandate | Governor | March 13 | March 16 | CAY | 54 | 509 | 132,872 |
| Arizona | Mandate | Governor and Department of Education | March 15 | March 16 | CAY | 670 | 2,320 | 1,110,834 |
| Arkansas | Mandate | Governor | March 15 | March 17 | CAY | 262 | 1,071 | 496,085 |
| California | Recommendation | Governor | March 19 | March 19 | REC | 1,027 | 10,248 | 6,220,070 |
| Colorado | Mandate | Governor | March 18 | March 23 | CAY | 185 | 1,894 | 910,280 |
| Connecticut | Mandate | Governor | March 15 | March 17 | CAY | 203 | 1,031 | 531,280 |
| Delaware | Mandate | Governor | March 13 | March 16 | CAY | 43 | 228 | 136,293 |
| District of Columbia | Mandate | Mayor | March 13 | March 16 | CAY | 62 | 224 | 87,308 |
| Florida | Recommendation | Department of Education/Superintendent | March 13 | March 16 | REC | 76 | 4,322 | 2,833,094 |
| Georgia | Mandate | Governor | March 16 | March 18 | CAY | 213 | 2,305 | 1,768,642 |
| Hawaii | Mandate | Department of Education/Superintendent | March 19 | March 23 | CAY | 1 | 292 | 180,837 |
| Idaho | Mandate | Department of Education/Superintendent | March 23 | March 24 | REC | 160 | 738 | 301,186 |
| Illinois | Mandate | Governor | March 13 | March 17 | CAY | 954 | 4,244 | 2,004,723 |
| Indiana | Mandate | Governor | March 19 | March 20 | CAY | 402 | 1,910 | 1,054,030 |
| Iowa | Mandate | Governor | March 15 | March 16 | CAY | 333 | 1,322 | 511,850 |
| Kansas | Mandate | Governor | March 17 | March 18 | CAY | 290 | 1,315 | 497,088 |
| Kentucky | Recommendation | Governor | March 12 | March 16 | REC | 176 | 1,533 | 680,978 |
| Louisiana | Mandate | Governor | March 13 | March 16 | CAY | 199 | 1,389 | 715,135 |
| Maine | Recommendation | Governor | March 15 | March 16 | REC | 223 | 585 | 180,204 |
| Maryland | Mandate | Department of<br>Education/Super<br>intendent | March 12 | March 16 | CAY | 25 | 1,420 | 893,684 |
| Massachusetts | Mandate | Governor | March 15 | March 17 | CAY | 406 | 1,849 | 954,031 |
| Michigan | Mandate | Governor | March 12 | March 16 | CAY | 886 | 3,720 | 1,515,370 |
| Minnesota | Mandate | Governor | March 15 | March 18 | CAY | 528 | 2,518 | 884,949 |
| Mississippi | Mandate | Governor | March 19 | March 19 | CAY | 157 | 1,060 | 478,321 |
| Missouri | Mandate | Governor and<br>Department of<br>Health | March 19 | March 19 | CAY | 560 | 2,387 | 915,412 |
| Montana | Mandate | Governor | March 15 | March 16 | VAR | 401 | 820 | 146,657 |
| Nebraska | Mandate | Governor | March 16 | March 23 | CAY | 249 | 1,052 | 323,766 |
| Nevada | Mandate | Governor | March 15 | March 16 | CAY | 20 | 707 | 485,785 |
| New Hampshire | Mandate | Governor | March 15 | March 16 | CAY | 191 | 493 | 179,554 |
| New Jersey | Mandate | Governor | March 16 | March 18 | CAY | 656 | 2,536 | 1,408,058 |
| New Mexico | Mandate | Governor | March 13 | March 16 | CAY | 150 | 871 | 334,341 |
| New York | Mandate | Governor | March 16 | March 18 | CAY | 1,005 | 4,699 | 2,724,663 |
| North Carolina | Mandate | Governor | March 14 | March 16 | CAY | 292 | 2,646 | 1,553,513 |
| North Dakota | Mandate | Governor | March 15 | March 16 | CAY | 173 | 469 | 109,823 |
| Ohio | Mandate | Governor and<br>Department of<br>Health | March 12 | March 17 | CAY | 960 | 3,539 | 1,704,399 |
| Oklahoma | Mandate | Department of<br>Education/Super<br>intendent | March 16 | March 17 | CAY | 543 | 1,800 | 695,092 |
| Oregon | Mandate | Governor | March 12 | March 16 | CAY | 202 | 1,247 | 580,645 |
| Pennsylvania | Mandate | Governor | March 13 | March 16 | CAY | 721 | 2,914 | 1,726,809 |
| Rhode Island | Mandate | Governor | March 13 | March 16 | CAY | 60 | 317 | 142,949 |
| South Carolina | Mandate | Governor | March 15 | March 16 | CAY | 88 | 1,244 | 777,507 |
| South Dakota | Recommendation | Governor | March 13 | March 16 | REC | 150 | 691 | 137,529 |
| Tennessee | Recommendation | Governor | March 16 | March 20 | REC | 147 | 1,782 | 1,001,967 |
| Texas | Mandate | Governor | March 19 | March 23 | CAY | 1,203 | 8,909 | 5,401,341 |
| Utah | Mandate | Governor | March 13 | March 16 | CAY | 154 | 1,050 | 668,274 |
| Vermont | Mandate | Governor | March 15 | March 18 | CAY | 212 | 309 | 87,769 |
| Virginia | Mandate | Governor | March 13 | March 16 | CAY | 134 | 2,015 | 1,291,462 |
| Washington | Mandate | Governor | March 13 | March 17 | CAY | 317 | 2,422 | 1,110,367 |
| West Virginia | Mandate | Governor | March 13 | March 16 | CAY | 55 | 706 | 272,266 |
| Wisconsin | Mandate | Department of Health | March 13 | March 18 | CAY | 445 | 2,257 | 860,752 |
| Wyoming | Mandate | Department of Health | March 19 | March 20 | VAR | 57 | 365 | 94,190 |
| Total | NA | NA | NA | NA | NA | 16,818 | 97,767 | 50,556,478 |

\*CAY, closed for academic year (through May or June 2020); REC, recommended to be closed for rest of academic year (through May or June 2020); VAR, varies by school/district.

Of the 50 states and DC, the governor or mayor (DC) issued the SC policy in 40 states and DC (80.4%); the state’s education authority (e.g. department or board of education, state superintendent) issued the policy in five states (9.8%); the state’s health authority (e.g. department of health, director of health, state health officer) issued the policy in two states (3.9%); and the governor and state health agency or state education authority issued the policy in three states (5.9%).

SCs were mandated in 44 states and DC and recommended in 6 states. The six states that recommended SCs were California, Florida, Kentucky, Maine, South Dakota, and Tennessee. Nebraska and Iowa initially recommended SC; however, schools were subsequently ordered to close on April 1, 2020 and April 2, 2020, respectively, approximately two weeks after the recommendation. Of the 44 states and DC where mandates were issued, schools in 41 states and DC were further required to remain closed for the rest of the academic year. Likewise, states that only recommended closure extended the recommendation through the end of the academic year.

Among states that mandated SCs, three states (Idaho, Montana, and Wyoming) did not order closure for the rest of the academic year. Idaho issued a mandated “soft closure” which gave schools the option of reopening before the academic year ended if they met specific state and local criteria. Despite the option of reopening before the end of the academic year in May or June 2020, no Idaho public school districts reported reopening. Montana and Wyoming schools could return to session at the discretion of local school boards and county health officials, respectively. Of the more than 400 districts in Montana, 14 districts (an estimated total of 16 schools serving less than 300 total students) reported a full reopening. All reopened districts in Montana were in rural locales, and all were in counties with no or very few new cases (<4) in the two weeks leading up to reopening [13]. Meanwhile, no Wyoming school districts reported a full reopening for traditional in-school learning.

While some state-level SC policies were issued as recommendations, to the best of our knowledge there was full compliance among public school districts. Therefore, because of prolonged statewide closures in response to COVID-19 transmission, more than 50 million public school students in nearly 100,000 schools from more than 16,000 school districts were impacted by COVID-related closures that lasted at least 8 weeks.

In addition to closing public K-12 schools, government policies in 19 states also specifically mandated the closure of private schools, and 4 states and DC later ordered closure of private schools via stay-at-home orders (SAH) (Fig 3). In these 23 states and DC, we estimated at least 9,700 private schools serving over 1,600,000 students were ordered to close in addition to public SCs.

**Figure 3.**
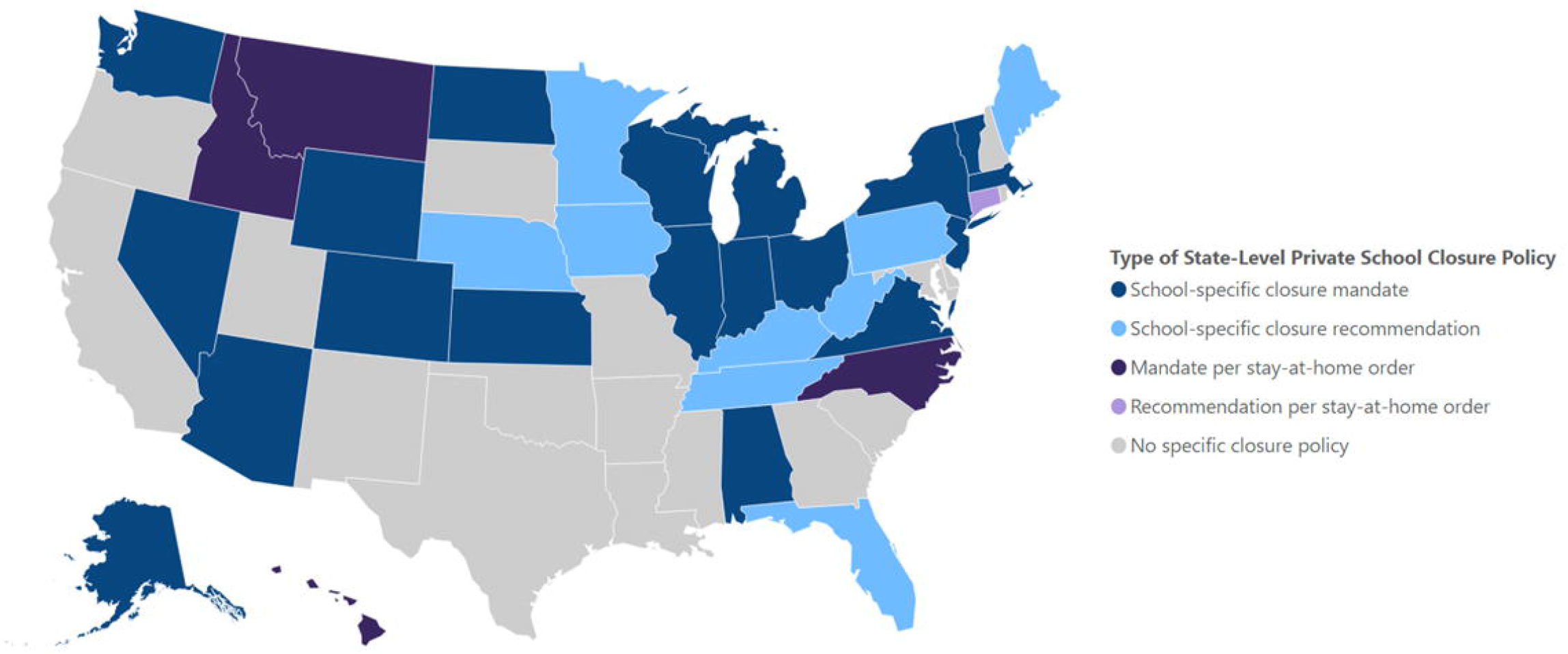
Type of Statewide Coronavirus Disease 2019 Private K-12 School Closure Policies-United States, March - June 2020. **Data Sources, Reference, & Notes:** Data were collected via daily searches of publicly available online content (Google Alerts, Google, and Google News). A standard set of search termswere used to identify COVID-19-related school closures lasting ≥1 day. Official state-level health and education department websites, and school and district level websites were also searched. The map shows the type of state-level closure policies for the academic year 2019-2020 for private K-12 schools in which schools are physically closed for students. Schools may have used distance learning to continue education from home. Orders and recommendations for statewide closure originated from state governors and/or state education departments.

### Continued provision of education and subsidized meals during COVID-19-associated closures

In response to prolonged statewide closures, school districts were faced with the need to mitigate the effects of secondary outcomes, namely the disruption of both learning and necessary subsidized meal programs.

Of the 600 school districts sampled, the majority [465 (77.5%)] offered continuity of education through distance learning for the duration of the closure; additionally, 81 (13.5%) districts offered distance learning for part of the closure, in which districts began offering distance learning and then suspended it at some time (Table 2). None of the districts sampled specifically announced that they would not offer distance learning, although for 54 (9%) sampled districts we could not ascertain whether distance learning was offered.

**Table 2.**
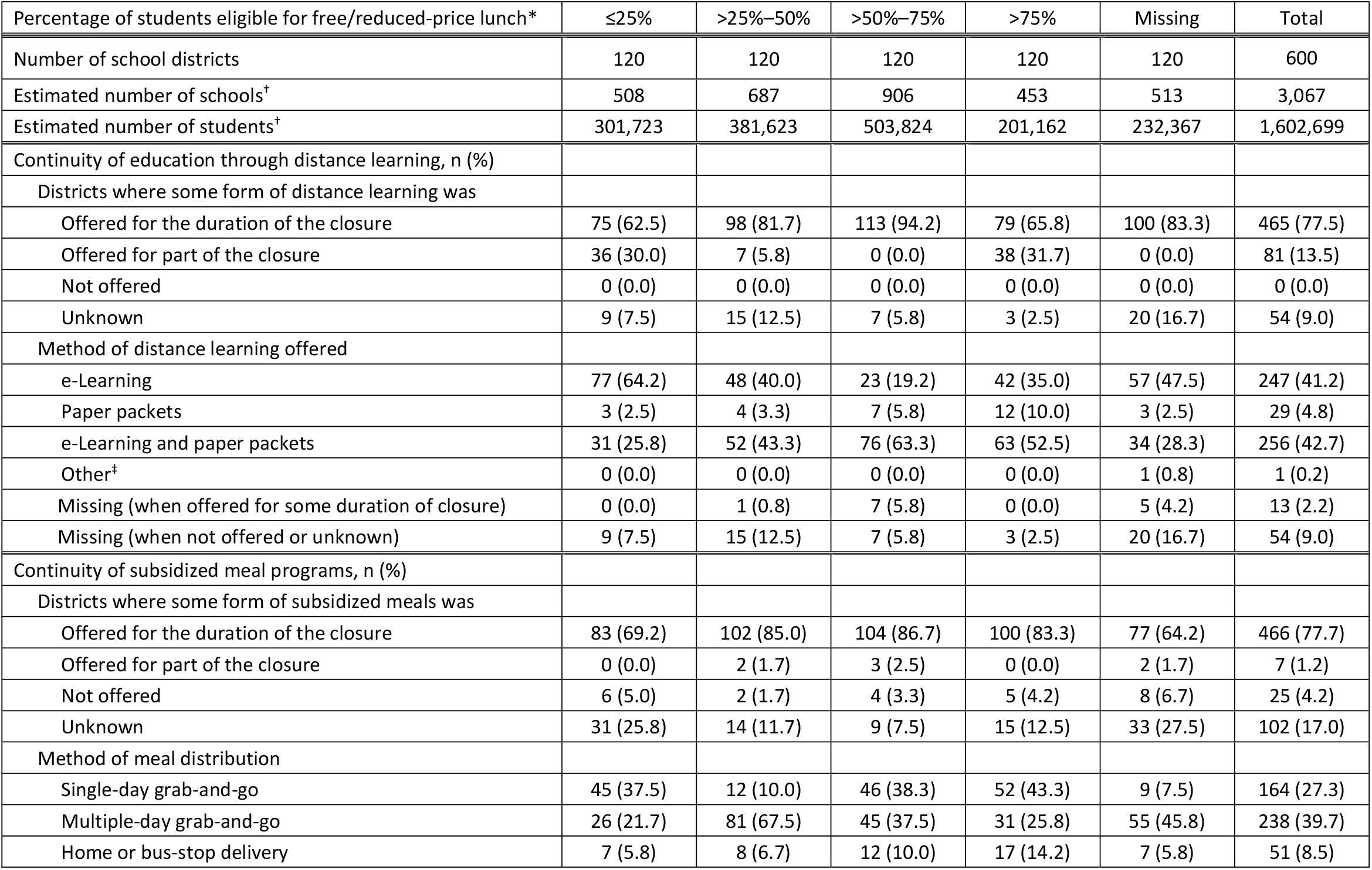
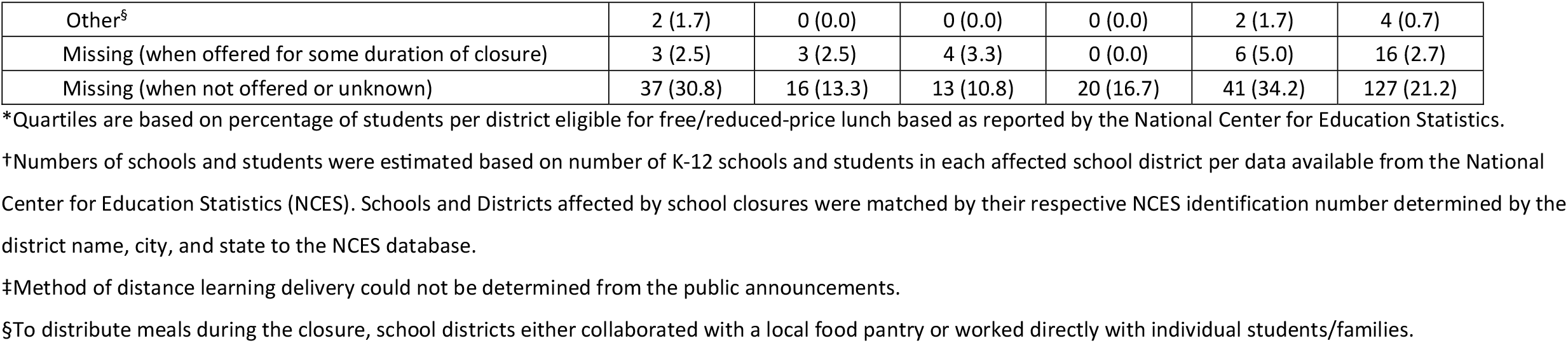
Characteristics of sampled school districts (N=600) describing the continuity of distance learning and subsidized meal programs during prolonged school closures due to the coronavirus disease 2019 pandemic — United States, March–May 2020.

| Percentage of students eligible for free/reduced-price lunch* | ≤25% | >25%–50% | >50%–75% | >75% | Missing | Total |
| --- | --- | --- | --- | --- | --- | --- |
| Number of school districts | 120 | 120 | 120 | 120 | 120 | 600 |
| Estimated number of schools<sup>†</sup> | 508 | 687 | 906 | 453 | 513 | 3,067 |
| Estimated number of students<sup>†</sup> | 301,723 | 381,623 | 503,824 | 201,162 | 232,367 | 1,602,699 |
| Continuity of education through distance learning, n (%) |  |  |  |  |  |  |
| Districts where some form of distance learning was |  |  |  |  |  |  |
| Offered for the duration of the closure | 75 (62.5) | 98 (81.7) | 113 (94.2) | 79 (65.8) | 100 (83.3) | 465 (77.5) |
| Offered for part of the closure | 36 (30.0) | 7 (5.8) | 0 (0.0) | 38 (31.7) | 0 (0.0) | 81 (13.5) |
| Not offered | 0 (0.0) | 0 (0.0) | 0 (0.0) | 0 (0.0) | 0 (0.0) | 0 (0.0) |
| Unknown | 9 (7.5) | 15 (12.5) | 7 (5.8) | 3 (2.5) | 20 (16.7) | 54 (9.0) |
| Method of distance learning offered |  |  |  |  |  |  |
| e-Learning | 77 (64.2) | 48 (40.0) | 23 (19.2) | 42 (35.0) | 57 (47.5) | 247 (41.2) |
| Paper packets | 3 (2.5) | 4 (3.3) | 7 (5.8) | 12 (10.0) | 3 (2.5) | 29 (4.8) |
| e-Learning and paper packets | 31 (25.8) | 52 (43.3) | 76 (63.3) | 63 (52.5) | 34 (28.3) | 256 (42.7) |
| Other<sup>‡</sup> | 0 (0.0) | 0 (0.0) | 0 (0.0) | 0 (0.0) | 1 (0.8) | 1 (0.2) |
| Missing (when offered for some duration of closure) | 0 (0.0) | 1 (0.8) | 7 (5.8) | 0 (0.0) | 5 (4.2) | 13 (2.2) |
| Missing (when not offered or unknown) | 9 (7.5) | 15 (12.5) | 7 (5.8) | 3 (2.5) | 20 (16.7) | 54 (9.0) |
| Continuity of subsidized meal programs, n (%) |  |  |  |  |  |  |
| Districts where some form of subsidized meals was |  |  |  |  |  |  |
| Offered for the duration of the closure | 83 (69.2) | 102 (85.0) | 104 (86.7) | 100 (83.3) | 77 (64.2) | 466 (77.7) |
| Offered for part of the closure | 0 (0.0) | 2 (1.7) | 3 (2.5) | 0 (0.0) | 2 (1.7) | 7 (1.2) |
| Not offered | 6 (5.0) | 2 (1.7) | 4 (3.3) | 5 (4.2) | 8 (6.7) | 25 (4.2) |
| Unknown | 31 (25.8) | 14 (11.7) | 9 (7.5) | 15 (12.5) | 33 (27.5) | 102 (17.0) |
| Method of meal distribution |  |  |  |  |  |  |
| Single-day grab-and-go | 45 (37.5) | 12 (10.0) | 46 (38.3) | 52 (43.3) | 9 (7.5) | 164 (27.3) |
| Multiple-day grab-and-go | 26 (21.7) | 81 (67.5) | 45 (37.5) | 31 (25.8) | 55 (45.8) | 238 (39.7) |
| Home or bus-stop delivery | 7 (5.8) | 8 (6.7) | 12 (10.0) | 17 (14.2) | 7 (5.8) | 51 (8.5) |
| Other<sup>§</sup> | 2 (1.7) | 0 (0.0) | 0 (0.0) | 0 (0.0) | 2 (1.7) | 4 (0.7) |
| Missing (when offered for some duration of closure) | 3 (2.5) | 3 (2.5) | 4 (3.3) | 0 (0.0) | 6 (5.0) | 16 (2.7) |
| Missing (when not offered or unknown) | 37 (30.8) | 16 (13.3) | 13 (10.8) | 20 (16.7) | 41 (34.2) | 127 (21.2) |

\*Quartiles are based on percentage of students per district eligible for free/reduced-price lunch based as reported by the National Center for Education Statistics.
†Numbers of schools and students were estimated based on number of K-12 schools and students in each affected school district per data available from the National Center for Education Statistics (NCES). Schools and Districts affected by school closures were matched by their respective NCES identification number determined by the district name, city, and state to the NCES database.
‡Method of distance learning delivery could not be determined from the public announcements.
§To distribute meals during the closure, school districts either collaborated with a local food pantry or worked directly with individual students/families.

Most of the sampled districts [466 (77.7%)] offered continuity of subsidized meal programs for the duration of the closure; an additional 7 (1.2%) offered subsidized meals for part of the closure, where meals were offered and then suspended at some time. Twenty-five (4.2%) districts did not offer subsidized meals, and we could not ascertain the status of meal programs for 102 (17.0%) of the districts in our sample. Of the districts that offered meals for all or part of the closure, the majority [402 (85.0%)] offered grab-and-go meals, 51 (10.8%) offered home or bus-stop delivery, and 4 (0.8%) offered other methods of meal distribution, whereby districts either collaborated with a local food pantry or worked directly with students or families (Table 2). The majority [21 (84.0%)] of districts that did not offer subsidized meals directed students and their families to alternative sources for meals (i.e. other district(s), community services).

We were able to ascertain specific reasons for 4 of the 7 districts which offered meals for only part of the closure. These four districts attributed their suspension of meal service either to their respective governor’s SAH or SC order, or to staff or student safety concerns because of COVID-19 (Table 3). Although public schools were not mandated by the federal government to provide meals, the US Department of Agriculture (USDA) took measures, in the form of national waivers, to ensure states had the flexibility to approve school provision of meals despite prolonged closure [14].

**Table 3.** School district-provided reasons when subsidized meal programs were offered for only part of the closure or not offered during the closure due to the coronavirus 2019 pandemic (N=25) – United States, March – May 2020.

|  | **Total** |
| --- | --- |
| Meals offered for part of the closure, n (%)<sup>*</sup> | 7 |
| Governor's school closure order<sup>†</sup> | 1 (14.3) |
| Governor's stay-at-home order<sup>†</sup> | 1 (14.3) |
| Safety concerns due to COVID-19 | 1 (14.3) |
| Governor's stay-at-home order and safety concerns due to COVID-19<sup>†</sup> | 1 (14.3) |
| Unspecified | 3 (42.9) |
| Meals not offered during closure, n (%) | 25 |
| Directed students to alternative sources (i.e. other district(s), community services) | 21 (84.0) |
| Made arrangements with families who faced hardship | 1 (4.0) |
| Safety concerns due to COVID-19 | 1 (4.0) |
| Sent checks to families who qualified for subsidized meals | 1 (4.0) |
| Unspecified | 1 (4.0) |

<sup>\*</sup>Two districts were unable to serve meals for 1-2 days due to environmental events (power outage, weather).
<sup>†</sup>The district decisions to suspend meal distribution in response to governor school closure or stay-at-home orders are not clear as the orders allowed for schools to prepare meals for eligible students.

## Discussion

### Magnitude of COVID-SCs

To our knowledge, the COVID-19–associated SCs that occurred during February–June 2020 represent the first ever, near-simultaneous implementation of school closure nationwide as an NPI in the US for any reason, including a pandemic. During this period, all of the nearly 100,000 public K-12 schools were concurrently closed for 8-13 weeks per policies implemented by their respective states, affecting more than 50 million public K-12 students. Hence, the COVID-SCs during the Spring semester of 2020 (February–June) were also the broadest ever recorded closures in duration and number of schools and students affected.

### SCs in historical context

Prior to the still ongoing COVID-19 pandemic, school closures in the US were broadly implemented only during the response to the 1918 influenza pandemic [15]. A modern-time retrospective study of NPIs used during the 1918 influenza pandemic found that schools across 43 cities were closed for a median duration of 6 weeks (range, 0–15 weeks) [16], often in combination with other mitigation tactics. However, unlike February–June 2020 when COVID-19–related closures were primarily implemented preemptively prior to widespread transmission, the closures in 1918 were asynchronous in their application across the country [17]. Moreover, unlike during the effectively nationwide closure of K-12 schools in response to COVID-19, in 1918 several large cities, including New York City and Chicago, formally kept their schools open throughout the pandemic [18]. Further, some cities—Philadelphia, for example—delayed implementation of mitigation measures including school closures until late into their local outbreaks. Comparative contemporary analyses showed that such delayed interventions were significantly less effective in reducing pandemic mortality than interventions implemented ahead of widespread virus transmission [16,19].

For pandemic influenza preparedness, only preemptive school closures—i.e. those closures implemented before illness becomes widespread among students and staff—are considered an effective NPI based on evidence from historic, epidemiologic, and modeling studies [1,20-23]. However, the evidence base on the effectiveness of reactive closures in reducing influenza transmission and delaying the epidemic peak remains divided with several studies reporting little or no impact on transmission [24-28] and others estimating reductions in transmission [29-31]. Of note, all of these studies were focused on the effects of reactive school closures during the seasonal or pandemic influenza outbreaks, and none evaluated the effects on SARS-CoV-2 transmission.

### Effectiveness of COVID-SCs

The effectiveness of the COVID-SCs will likely be a topic of much research for years to come. Several studies completed so far suggest that SCs implemented during the initial six months of the COVID-19 pandemic (January–June 2020) in the US and elsewhere resulted in reduced transmission and mortality. A study analyzing COVID-SCs in the US during March 9–May 7, 2020 concluded that SCs were temporally associated with significant decreases in both incidence of and mortality attributed to COVID-19; states that closed schools when cumulative incidence was lower experienced greater reductions in incidence and mortality than states that closed schools later in their relative epidemic curves [32]. These findings were congruent with another US study, which found that higher mortality was associated with the later statewide SCs in relation to the date of the first COVID-19–related death in the state [33]. The impact of COVID-SCs might also have been bolstered by the additional 2–3-month break from classroom congregation during the subsequent summer recess. Further, in a study evaluating the impact of COVID-19 NPIs on a global scale, COVID-SCs along with restrictions of mass gatherings and implementation of physical distancing showed a strong negative association with epidemic growth [34]. However, addressing the question of effectiveness will require much more additional study over the course of this ongoing pandemic.

### Continuity of education and services during COVID-SCs

The wave of COVID-19–associated SCs during the second semester of the 2019–2020 school year (February–June 2020) was followed by the largest ever nearly simultaneous transition to various types of distance learning nationwide in lieu of congregating students and staff at schools. Responding to the unprecedented disruption to normal functions, US school districts swiftly mobilized distance learning programs and alternative feeding programs to preserve the continuity of education and services on which students and their families rely as recently noted by others [35]. Prior research supports the importance of continuity of education and the subsidized meal programs. Research on gaps in learning, such as during summer breaks, suggests that learning loss might occur during extended periods away from traditional, in-person instruction [36]. Similarly, studies have also reported reductions in expected achievement during weather-related closures [37] or in relation to increased student absenteeism [38]. Furthermore, schools are also providers of ancillary services on which students and their families rely. Previous research indicates that school-provided meals, such as those via the National School Lunch Program, are associated with decreased food insecurity, improved nutritional health, and increased academic performance [39,40]. The continuity of education and other school-based services likely serves to mitigate the disruption caused by the sudden and prolonged nature of SCs during the initial five months of the COVID-19 pandemic [40].

Among a sample of 600 public K-12 districts, the majority offered distance learning and continued provision of subsidized meal programs (546 [91%] and 474 [79%], respectively) during the closures. These high rates of substitution for normal in-school education delivery and routine school-based meals occurred despite of the fact that, prior to the COVID-19 pandemic, less than 50% of school districts had plans in place for ensuring continuity of education or feeding of students during closure [41]. Additionally, while these services were frequently offered, further study could assess what proportion of students were consistently and effectively able to receive them.

### Challenges posed by COVID-SCs

Despite the efforts of school districts to ensure continuity of education and subsidized meal programs in the face of a deadly pandemic, the sudden, prolonged transition to out-of-school learning was not easy for many students, parents, or educators. According to national Gallup polls, 42% of parents were concerned about negative effects to their child’s education during COVID-SCs [42] and nearly three in ten parents said their child was suffering harm because of physical distancing [43]. Similarly, in nationally representative, biweekly polling of about 1,900 educators, the majority of educators reported both student and teacher morale to be either somewhat lower (42% and 40%, respectively) or much lower (16% and 31%, respectively) than prior to COVID-19. They also consistently reported lower engagement from students [44].

Research on pandemic preparedness in the education sector conducted prior to the 2020 COVID-19 pandemic identified opportunities and barriers to transitioning to distance learning compared to traditional school closure without educational activities [45,46]. Analyses of a large study among school practitioners (i.e. teachers, principals, superintendents) across the US identified barriers to implementing distance learning during prolonged SCs in response to emergencies [46]. Participants reported significant challenges for schools that had not implemented some form of online learning before an emergency to create online content, train staff, and provide access to technology and high-speed internet immediately after an emergency [45]. A review of both peer-reviewed literature and state-level pandemic planning for K-12 schools indicated that there were limited resources available to schools to inform the development of physical distancing policies and procedures [46].

In coping with this transition to distance learning during COVID-SCs, numerous reports and polls from across the US identified barriers to implementation among students and their families. These barriers included inadequate access to digital devices and reliable internet at home, lack of support for non-native English speakers, inequity in access to meals, and issues stemming from implementing new online learning strategies [46-50].

### Disparities and COVID-SCs

The barriers faced by students and families during the implementation of SCs in response to a pandemic could have been exacerbated among racial/ethnic minority populations and those of lower socioeconomic status because of pre-existing disparities, which impact access to education and food security [51-53]. The study of racial/ethnic disparities in the context of SCs predates the current pandemic and was documented a year before the markedly less severe 2009 Influenza A (H1N1) pandemic [54]. As described during a CDC stakeholder meeting convened to assess these barriers, the major challenges to racial/ethnic minority populations associated with SCs include lack of free lunches, unmet educational needs because of the disruption of traditional in-school learning, and lack of childcare options while parents are away at work [54]. Stakeholders identified reasons why these populations would encounter barriers to adopting interventions, which include, but are not limited to, socioeconomic factors and lack of educational materials and communications tailored to culturally and linguistically diverse populations.

These findings on the disproportionate impacts of SCs are reinforced by the results of surveys completed by parents in four states (California, New York, Texas, Washington) during March and April 2020, which indicated that parents, especially those from African American, Hispanic, and low-income families, were concerned that their children were falling behind academically and reported gaps in access to both digital resources and meals during COVID-SCs [50]. These burdens were compounded by disproportionate incidence of COVID-19 among racial/ethnic minorities, whereby these populations encountered both greater risk of the disease and lesser access to safer forms of instruction [55].

Despite the negative impact of COVID-SCs, several statewide polls indicated that voter/parental support for extended closures during the initial months of the COVID-19 pandemic was high (≥70% in all polls, [56-60]; and >90% in one poll, [56]). In a national Gallup Panel Poll conducted in March and April 2020, the majority of parents (56%) preferred children to have in-school learning full-time in the coming 2020╌2021 school year, with a greater proportion of working parents (59%) expressing this preference than non-working parents (46%), however the results were not presented by race/ethnicity [61]. At the same time, less than half (46%) of Americans supported returning to school prior to availability of a vaccine [62].

## Limitations

Our results should be considered in conjunction with at least six limitations. First, the data collected through daily online searches might not be comprehensive. This might be attributed to the hierarchy of results produced by search engines, some news not being reported to mainstream sources, and local news from small, rural, or independent news sources potentially less likely to be captured by searches. Further, because data were abstracted using information available in publicly available closure announcements, some information might not have been complete or entirely accurate. However, this study benefited from the existence of an established monitoring system used to routinely collect data on influenza-related SCs [10,63], which was quickly modified to capture COVID-SCs in near real time. Data facilitated daily updates to the CDC COVID-19 Response Incident Manager from February 27, 2020, when COVID-SCs began and then gradually progressed into the nationwide closure. Second, district closure data might not have been publicly available and captured by our daily searches if only sent directly to families (via email, text, phone call, etc.). We mitigated this possible limitation by also acquiring the data from other publicly available sources such as official school and school district websites, Facebook pages, and Twitter feeds, which did not rely on direct reporting requirements from school districts. Third, the duration of statewide closures is likely underestimated for most affected schools because we do not know the specific date that each public school district was expected to begin their summer recess. The range of dates for the start of summer break are understood to be from mid-May until late June; therefore, this date was conservatively estimated to be at the completion of the third week of May. We also chose to only estimate the number of weeks rather than student-days or hours lost because of the variation across school calendars that could make those figures less reliable. Fourth, we could have overestimated the number of districts that had continuity of education and subsidized meal programs throughout the closure because, unless a district expressly stated distance learning and/or subsidized meals was offered for only part of the closure, we assumed they were offered for the entire duration of closure. Fifth, we were unable to find information for continuity of distance learning for 9% of districts and for subsidized meal programs for 17% of districts; however, information was complete for the majority (>80%) of the sampled districts. Finally, the sample data collection on how districts managed the continued provision of education and meals throughout prolonged SC was not intended to be representative of all U.S. districts, but rather to give insight into the ways schools across the U.S. were able to cope. Nonetheless, by stratifying the sample selection according to percentage of students eligible for free and reduced-price lunch, we included school districts across a spectrum of economic strata. Additionally, the school districts in the sample spanned US geography, including all ten HHS regions and 46 of 50 states.

## Conclusion

In conclusion, the initial introduction of SARS-CoV2 into the US was followed by the unprecedented, nearly simultaneous nationwide closure of all public K-12 schools, the majority of which transitioned to distance learning, affecting more than 50 million students. Despite this, schools, staff, and students across the nation demonstrated flexibility as they coped with the disruption of traditional instruction and services. However, reported disparities in resources for continued education and subsidized meal programs are troubling, especially because of the apparent overlap with the greater COVID-19 burden among the same racial/ethnic minority groups. The disproportionate impact to students and families from groups at higher risk for severe outcomes from COVID-19 should be further studied and, where needed, remedied as communities continue to experience COVID-19 outbreaks.

## Data Availability

The authors confirm that all data underlying the findings are fully available without restriction. Data are available from Google (www.google.com), Google News (news.google.com), and the National Center for Education Statistics (nces.ed.gov). The search and linking strategy used for these data sources is detailed within the paper.

## Acknowledgments

The authors would like to thank the team who assisted with the monitoring of COVID-19-associated school closures, including Ashley Jackson, Cassandra Kersten, Peter Kim, and Darielle Oliver. The authors would also like to thank Meeyoung Park and Sarah Moreland for their contributions in generating maps and Faruque Ahmed for his guidance regarding edits to data collection forms to include COVID-19-specific variables.

